# Anxiolytic effects of a galacto-oligosaccharides prebiotic in healthy female volunteers are associated with reduced negative bias and the gut bacterial composition

**DOI:** 10.1101/19011403

**Authors:** Nicola Johnstone, Chiara Milesi, Olivia Burn, Bartholomeus van den Bogert, Arjen Nauta, Kathryn Hart, Paul Sowden, Philip WJ Burnet, Kathrin Cohen Kadosh

## Abstract

Current research implicates pre- and probiotic supplementation as potential mediators for improving symptomology in numerous physical and emotional ailments. The alteration of emotional states via nutrient intake is an attractive concept for clinicians and consumers alike and may be an efficient channel to improved well-being. Here we focus on the period of late adolescence, which is a time of emotional refinement via maturation that significantly influences emotional, social and physical well-being in later years. Effective interventions such as nutritional supplementation in this age group have the potential to offset health-related costs in later life. In this study we examined multiple indices of mood and well-being in 64 healthy females in late adolescence in a 4-week double blind, placebo controlled Galacto-oligosaccharide (GOS) prebiotic supplement intervention. We also obtained stool samples at baseline and follow-up for microbiome sequencing and analyses. We found effects of the GOS intervention on sub-clinical self-reported high trait anxiety, attentional bias, and bacterial abundance, suggesting that dietary supplementation with a GOS prebiotic may be influential in improving indices of pre-clinical anxiety.

## Introduction

In recent years, the gut microbiome has emerged as an important player in our efforts to understand the factors that influence brain function and behaviour^1–4^. The gut and the brain are intimately connected via the gut-brain axis, which involves bidirectional communication via neural, endocrine and immune pathways^5–7^. For example, gut microbial composition — which itself alters throughout the lifespan and in response to factors such as stress and lifestyle choices including diet^8^ — has been shown to regulate gene expression and the release of metabolites in the brain^9,10^. There are also suggestions that a significant reduction in microbial diversity, or an increased number of pathogenic microbes, affects brain-behaviour relationships, and may lead to psychological abnormalities that underlie mental illness^11^. To date, studies in humans have focused on characterising microbe populations in both health and disease^4,12^. In adults for example, the gut microbiota has been shown to be related to atypical social functioning in autism^13^ and symptoms of anxiety and depression^7,14^. Animal research has suggested that the gut microbiota plays an important role during key moments in the host development, and in particular during adolescence which represents a critical time window where microbiota help fine-tune the gut-brain axis^3,8,15^. One of the consequences gut microbiota variations during neurodevelopment is that it may lead to aberrant brain network maturation and thus atypical behavioural patterns. This supposition highlights the importance of understanding how changes in the gut microbiome relate to brain function and plasticity during this critical developmental period^16^.

Animal research has highlighted the significant impact of the gut microbiota on the development and maturation of brain networks that underlie emotion^15–17^. Particularly, nutritional interventions have been shown to fortify the microbiome-gut-brain axis and ameliorate microbial imbalances such as dysbiosis. Drastic changes in diet can alter microbial diversity in a matter of days^18^, and research also suggests that modifying microbial ecology via the intake of so-called ‘*psychobiotics*’ could help reduce stress responses and symptoms of anxiety and depression^19–21^. The term psychobiotics refers to live cultures of beneficial gut bacteria (*probiotics*) or substrates which enhance the growth and/or activity of indigenous beneficial intestinal bacteria (prebiotics), which when ingested in sufficient amounts improve brain function^22,23^. Probiotic strains, including members of the genera *Lactobacillus* and *Bifidobacterium*, are enriched in some dairy/fermented products, whereas prebiotics are non-digestible substances that feed the gut microbiome^18,24^ such as fructans and oligosaccharides found in in cereals, fruits and vegetables^22^. Both pro- and prebiotic substances are commercially available as supplements. There is no doubt that the administration of psychobiotics (both probiotics and prebiotics) to rodents leads to robust, reproducible, attenuating effects on anxious and depressive-like behaviours, and suppress the neuroendocrine stress response. However, the translatability of these psychotropic actions to humans remains unclear.

A recent study in adult volunteers found that the consumption of probiotics in a randomised double-blind trial, reduced measures of low mood and distress, and urinary free cortisol which indicated a decreased stress response^25^. Similarly, a 4-week course of a multispecies probiotic in healthy participants resulted in reduced responsiveness to sad mood^26^. A neuroimaging investigation in female participants who had consumed a probiotic containing yoghurt over 4 weeks, revealed differential signals over brain regions involved in emotion processing and regulation^27^. The main problem of using probiotics is that introducing an allochthonous probiotics species could disturb the cross-feeding microbiota population, which is particularly disruptive in participants with weakened immune systems^28^. Prebiotics on the other hand support the beneficial bacteria that are already present in the participants’ gut, such as *Bifidobacterium* which have been linked to emotional well-being^25–27^.

Intake of a galacto-oligosaccharides prebiotic over 4 weeks has also been shown to lower the secretion of the stress hormone cortisol and emotional processing in healthy adults^29^ in comparison to placebo. In the same study, participants exhibited decreased attentional vigilance to negative information in a dot-probe task. Given that anxious people routinely exhibit increased biases towards negative information^30^, this suggests that GOS intake may be useful in modifying anxiety-related psychological mechanisms.

The main aim of the current study was to investigate whether GOS intake influences anxiety and mood measures in late adolescence and early adulthood in humans. Specifically, we used emotion regulation as a model for anxiety, as good emotion processing abilities in development are linked with various indices of well-being and mental health^31^. Moreover, it has been repeatedly shown that adolescence represents an important developmental juncture for both the emergence of social fears^31^ and the development of emotion control abilities, which allow the individual to control their fear responses and anxiety^32^. Here we compared the effects of a 4-week course of GOS supplements compared to a placebo on the gut-microbiome and emotional behaviour and well-being. Emotional behaviour was assessed with the attentional dot-probe task^29^ and a number of self-report measures of anxiety^33,34^, depression^35,36^ and emotion regulation abilities^37,38^. In the current study we hypothesised that daily intake of galacto-oligosaccharides (GOS) for 4 weeks would: 1) reduce self-reported levels of anxiety in the GOS group in comparison to the placebo group; 2) improve attentional bias towards positive emotional stimuli in the dot-probe task, and 3) stimulate the fecal abundance of potently beneficial gut bacteria (such as *Bifidobacterium*) in the GOS compared to the placebo group.

## Materials and Methods

### Participants

Sixty-four healthy late adolescent female volunteers were recruited to this double-blind placebo-controlled 4-week galacto-oligosaccharides (GOS) supplement intervention study via posters and online advertisements. Inclusion criteria were no current or previous clinical diagnoses of anxiety or co-morbid neurological, psychiatric, gastrointestinal, or endocrine disorders; no current habitual use of prebiotic or probiotic supplements; no antibiotic use in the 3 months prior to study enrolment and no vegan diets. All participants provided written informed consent prior to testing and received financial compensation for participating in the study.

Participants were screened for trait anxiety scores on the STAI-Trait subscale^33^ and randomised with stratification for baseline anxiety to either GOS or placebo group based on anxiety level (high or low, median split of sample scores at baseline). This study was given a favourable opinion by the University of Surrey Ethics Committee (UEC/2017/086/FHMS), and is registered on https://www.clinicaltrials.gov number NCT03835468.

### Design

Power to detect statistically significant effects in behavioural measures was determined to be sufficient with a sample size of 60 participants, posited on small effect sizes found in similar research^29^. At both testing time points, baseline and follow-up, the same testing protocol was used. Specifically, all participants completed a comprehensive battery of self-report questionnaires (see materials for a description) assessing indices of anxiety^33,34^, mood^35,36^, emotion regulation^37,38^ and sleep^39^. The self-reported trait anxiety scores at baseline were used to group participants as high or low anxious based on the median score of the sample collected. Participants also completed two subscales (verbal and matrix reasoning) of the Wechsler Abbreviated Scale of Intelligence^40^ to estimate IQ, as well as the attentional dot-probe task to assess overt emotional processing^29^. Each participant was also provided with a stool-sampling kit (MyMicroZoo, Leiden, The Netherlands) to self-collect faeces for gut microbiome sequencing analysis. Following the baseline assessment, participants were issued a 28-day supply of supplements (GOS or placebo), which were to be taken once daily. To assess usual intake at the start of the study and to monitor compliance with the dietary instructions (to consume the supplement and to not change their usual diet) participants were also asked to complete a 4-day food estimated diary at each time point which was reviewed by a member of the research team at their testing appointments. The food diary is a semi-quantitative diary used routinely by our group, which required participants to record their intake concurrently during the day in as much detail as possible, including brand names, cooking methods and portion sizes. Diaries were analysed using nutritional analysis software (Nutritics.com) for energy and macro- and micronutrient intakes.

At follow-up, the entire testing protocol was repeated for each participant following cessation of supplement supply (study duration Mdn = 30 days for both GOS group (range: 25-36 days) and placebo group (range: 27 - 43 days)), and participants provided a second stool sample.

### Self-report measures

Participants completed a demographic questionnaire obtaining information on age, height, weight and relevant medical history. Following this, psychological self-assessment questionnaires obtained indices of anxiety, mood and emotion regulation. Anxiety assessments comprised measures of state and trait anxiety (State-Trait Anxiety Inventory; STAI^33^) and social anxiety (Social Anxiety Scale for Adolescents and Young People; SAS-A^34^). Mood measures included measures of general mood (Mood and Feelings Questionnaire: Short Version; MFQ^36^) and depression (Beck Depression Inventory-II; BDI-II^35^). Emotion regulation was indexed using the Emotion Regulation Questionnaire for Children and Adolescents (ERQ-CA^37^) and Thought Control Ability Questionnaire (TCAQ^38^). Finally, participants reported sleep quality for the preceding 1-month period using the Pittsburgh Sleep Quality index (PSQI^39^).

### Attentional dot probe task

The attentional dot-probe task was adapted from Schmidt and colleagues^29^ and presented E-Prime 3.0 software (Psychology Software Tools, Pittsburgh, PA) to assess emotional processing and biases to negative, positive and neutral word pairs in masked and unmasked conditions. Negative and positive words induce emotional bias, whereas neutral word words pairs act as fillers. Masked and unmasked conditions alter awareness of stimulus emotional valence. At the beginning of each trial, a fixation cross appeared on screen for 500 ms. This was followed with a word pair that was one of 30 positive-neutral, 30 negative-neutral and 30 neutral-neutral pairs, with words positioned at the top and bottom of the screen. Emotional word position was equally split across top and bottom. In the masked condition (90 trials), word pairs were presented for 17 ms, followed by a length and position matched mask of a nonsense letter string for 483 ms. In the unmasked condition (90 trials), word pairs were presented for 500 ms. This was followed by a probe of either one or two stars, that was congruent or incongruent with the emotional word position. The participant responded by pressing either ‘1’ or ‘2’ corresponding to the number of stars in the probe, terminating the trial. Frequency of number of stars and of position was equally split across all trials. A total of 180 trials were presented and randomised across masked and unmasked conditions, and valence stimulus pairs. Attentional vigilance was calculated from the response times of correct responses by subtracting congruent RT from incongruent RTs for positive and negative stimuli in masked and unmasked conditions separately. E-Prime 2.0 (Psychology Software Tools, Pittsburgh, PA) was used to present experimental stimuli.

### GOS/placebo supplement

Participants received either a daily dose of 7 grams of the prebiotic galacto-oliogosaccharides (GOS) provided by FrieslandCampina, Amersfoort, The Netherlands; or a placebo (maltodextrin, dried glucose syrup) for a period of 28 days. GOS are non-digestible carbohydrates, which are not completely broken down by human digestive enzymes. Because of this, they reach the intestine relatively intact, where they are then available for the present microbiota^23^ whereas Maltodextrin is absorbed in the upper part of the intestine and does not reach the colon. Both supplements were provided in powdered form in unlabelled packaging and are similar in colour and taste. Supplements were instructed to be consumed by mixing with food or drink once daily.

### Stool sampling

At baseline and follow-up participants were provided with a unique sampling kit provided by MyMicroZoo (Leiden, the Netherlands) for stool collection at home. Feces samples were collected in DNA/RNA Shield (Zymo Research, CA, USA) and returned by the subjects to the recruitment station where collected samples were returned and stored at −80°C prior to being shipped on dry-ice for analysis by MyMicroZoo.

### DNA extraction

DNA extraction was performed using the Quick-DNA(tm) Fecal/Soil Microbe Miniprep Kit (Zymo Research) according to manufacturer’s instructions except for using the fecal slurry, containing DNA/RNA Shield, as input during bead beating for mechanical cell lysis instead of using the lysis buffer provided in the extraction kit.

16S rRNA gene based bacterial profiling. Illumina 16S rRNA gene amplicon libraries were generated and sequenced at BaseClear (Leiden, the Netherlands). In short, barcoded amplicons from the V3-V4 region of 16S rRNA genes were generated using a 2-step PCR. 10 genomic (g)DNA was used as template for the first PCR with a total volume of 50 ul using the 341F (5’-CCTACGGGNGGCWGCAG-3’) and the 785R (5’-GACTACHVGGGTATCTAATCC-3’) primers appended with Illumina adaptor sequences. PCR products were purified, and the size of the PCR products were checked on Fragmentanalyzer (Advanced Analytical, CA, USA) and quantified by fluorometric analysis. Purified PCR products were used for the 2nd PCR in combination with sample-specific barcoded primers (Nextera XT index kit; Illumina, CA, Illumina). Subsequently, PCR products were purified, checked on a Fragment analyser and quantified, followed by multiplexing, clustering, and sequencing on an Illumina MiSeq with the paired-end (2x) 300 bp protocol and indexing. The sequencing run was analysed with the Illumina CASAVA pipeline (v1.8.3) with demultiplexing based on sample-specific barcodes. The raw sequencing data produced was processed removing the sequence reads of too low quality (only “passing filter” reads were selected) and discarding reads containing adaptor sequences or PhiX control with an in-house filtering protocol. A quality assessment on the remaining reads was performed using the FASTQC quality control tool version 0.10.0. The Illumina paired reads were merged into single reads (so-called pseudoreads) through sequence overlap, after removal of the forward and reverse primers. Chimeric pseudoreads were removed using USEARCH 9.2^41^ and the remaining reads were aligned to the RDP 16S rRNA gene database^42^. Based on the alignment scores of the pseudoreads, the taxonomic depth of the lineage is based on the identity threshold of the rank; Species 99%, Genus 97%, Family 95%, Order 90%, Class 85%, Phylum 80%. Illumina reads were deposited in the European Nucleotide Archive (ENA) database (http://www.ebi.ac.uk/ena) in fastq-format under study accession number PRJEB32693 (or secondary accession number ERP115404). Abundance of each genera was calculated as a percentage of the total number of sequences identified in each sample. Shannon entropy of counts, a metric of microbiota diversity, was calculated using USEARCH 9.2^41^ with OTU clustering with a sequencing identity threshold of 97% after subsampling from the entire set, to account for different sampling depths.

### Multivariate analysis

Partial Redundancy analysis (RDA) was performed using the rda-function in the Vegan-package (version 2.5-4^43^) in R (version 3.3.1) to assess correlations between 16S rRNA gene based bacterial composition data (at the genus level) and the GOS treatment at baseline versus follow-up (as environmental variable), after conditioning the data for the covariate ‘subject’. RDA can be considered a constrained version of Principal Component Analysis (PCA). Where PCA considers all variance encompassed in the data, RDA only considers variance explained by the environmental variables (in this case time, contrasting baseline versus follow-up). The psychological response variables SAS, STAI Trait, STAI state, MFQ, BDI from the self-report measures, and positive and negative bias from attentional vigilance calculations in the dot probe task were fitted onto the ordination. In the resulting biplot, a type 1 (object focused) scaling is employed so that the scores for samples are scaled by eigenvalues and that when plotted, the distances between them represent their similarity (Euclidean distances).

## Statistical analysis

Self-report measures obtained at follow-up were compared using analysis of covariance (ANCOVAs) between intervention groups (GOS and placebo) and anxiety grouping (High and Low) considering baseline measures as a covariate. Baseline measures were initially tested for differences between intervention groups prior to using ANCOVAs. Similarly, attentional dot-probe outcomes at follow-up were assessed for attentional vigilance to positive and negative stimuli in the masked and unmasked conditions using ANCOVAs between intervention and anxiety groups with baseline measures as a covariate. Bacterial diversity at follow up was also evaluated with ANCOVA examining intervention and anxiety groups. Bacterial abundance of all genera was evaluated using non-parametric calculations comparing intervention groups at baseline and follow-up (Wilcoxon rank-sum test for independent groups), and each intervention group across time (Wilcoxon signed rank test for paired samples). Unrestricted Permutation Test was used to assess significance of the GOS treatment and was indicated in the RDA diagrams. Statistical testing was performed in R (version 3.5.1^44^).

## Results

A 25% attrition rate resulted in 48 participants completing the study, n = 23 in the GOS group (high anxiety n = 12, low anxiety n = 11) and n = 25 in the placebo group (high anxiety n = 13, low anxiety n = 12), matched for age (GOS median (Mdn) = 19 years (range: 18-25), placebo Mdn = 20 years (range: 18-25)), IQ (GOS Mdn = 103 (83-148), placebo Mdn = 102 (72-125)) and BMI (GOS Mdn = 20.5 kg/m^2^ (range: 18.8-30.1), placebo = Mdn 20.3 kg/m^2^ (range: 16.2 – 27.2)). Diets were consistent across time for each supplement group for total energy consumption, although the GOS group had lower overall energy intake than placebo at both baseline (*p* = .009) and follow up (*p* = .025). At the macro level, consumption was estimated as a percentage of energy intake for each individual over the first (baseline) and last (follow up) four-day period of supplement intake. Between GOS and placebo groups, non-parametric tests showed no significant difference for carbohydrate, fat, protein, dietary saturated fat, sugars or alcohol intake. From baseline to follow up paired sample non-parametric tests showed no difference in any of the aforementioned measures in the placebo group, but the GOS group demonstrated a reduction in sugar consumption at follow up (*p* = .010), with all other measures remaining consistent (Supplementary table 1).

### Self-report measures

For all measures, models comparing baseline scores confirmed no differences between groups thus ANCOVA was used to compare group means at follow-up (**Table 1**). The significant interaction of intervention and anxiety grouping on trait anxiety scores *F*(1,42) = 5.58, *p* = .023, n^2^ = .03 was followed up with further ANCOVAs examining the influence of intervention for anxiety grouping independently. It was found that compared to the placebo group, four weeks of GOS consumption reduced self-reported scores for high anxious participants at trend-level *F*(1,21) = 3.88, *p* = .062, n^2^ = .12; adjusted means GOS *M* = 45.47, *SE* = 1.43 [CI 42.49 – 48.43]; placebo *M* = 49.45, *SE* = 1.43 [CI 46.48 – 52.42] but not for the low anxious group *F*(1,20) = 1.84, *p* = .190, GOS *M* = 32.34, *SE* = 1.59 [CI 29.03 – 35.65]; Placebo *M* = 29.36, *SE* = 1.52 [CI 26.19 – 32.52] (**Figure 1**). There were no other interactions between intervention group and anxiety group for social or state anxiety, or in mood measures (BDI or MFQ), emotion regulation or sleep quality index (**Supplementary Table 2**).

**Table 1.**
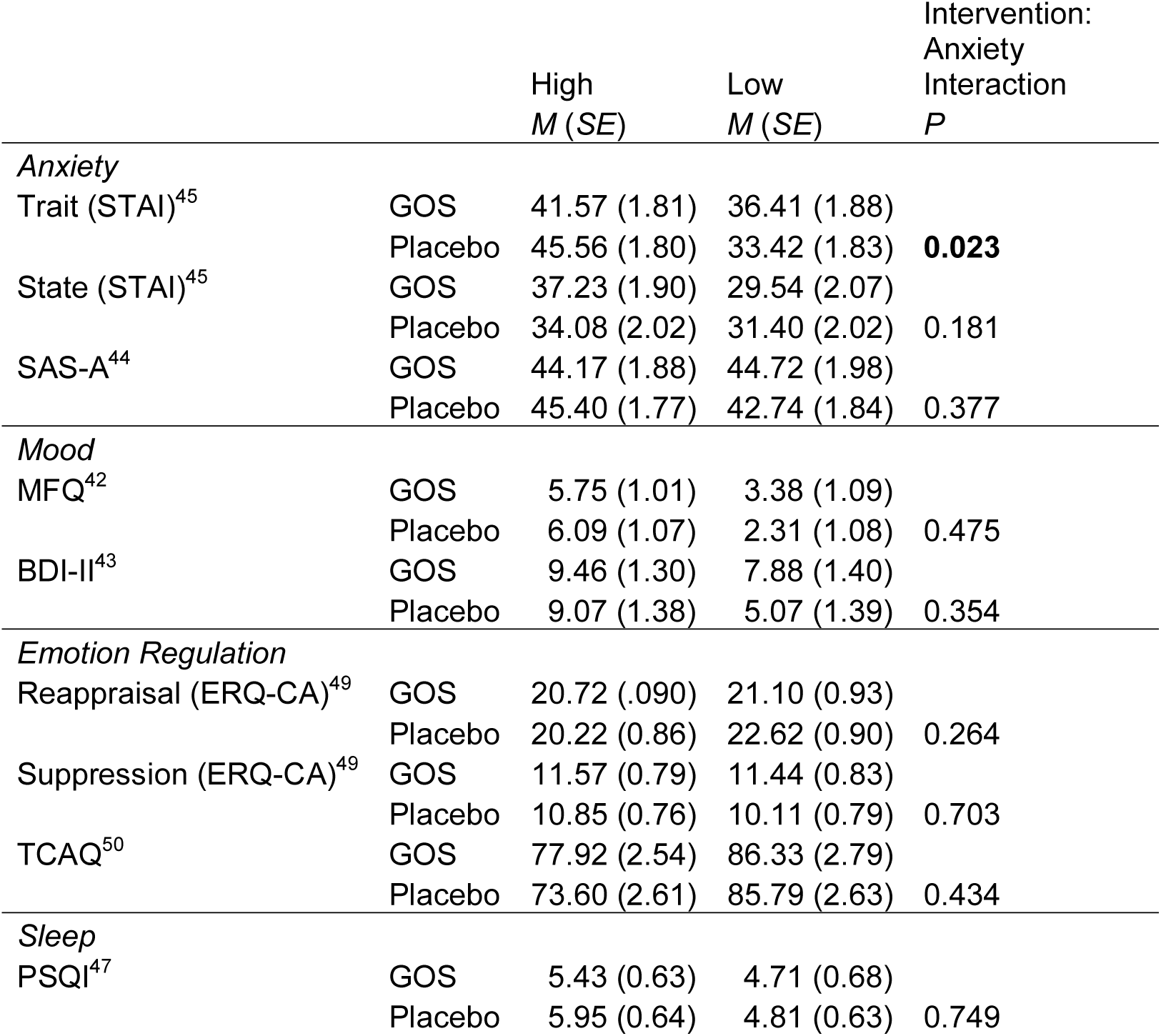
Self-report measure adjusted means at follow-up with ANCOVA interaction.

|  |  | High<br>*M (SE)* | Low<br>*M (SE)* | Intervention:<br>Anxiety<br>Interaction<br>*P* |
| --- | --- | --- | --- | --- |
| *Anxiety* |  |  |  |  |
| Trait (STAI)<sup>45</sup> | GOS | 41.57 (1.81) | 36.41 (1.88) | **0.023** |
|  | Placebo | 45.56 (1.80) | 33.42 (1.83) |  |
| State (STAI)<sup>45</sup> | GOS | 37.23 (1.90) | 29.54 (2.07) | 0.181 |
|  | Placebo | 34.08 (2.02) | 31.40 (2.02) |  |
| SAS-A<sup>44</sup> | GOS | 44.17 (1.88) | 44.72 (1.98) | 0.377 |
|  | Placebo | 45.40 (1.77) | 42.74 (1.84) |  |
| *Mood* |  |  |  |  |
| MFQ<sup>42</sup> | GOS | 5.75 (1.01) | 3.38 (1.09) | 0.475 |
|  | Placebo | 6.09 (1.07) | 2.31 (1.08) |  |
| BDI-II<sup>43</sup> | GOS | 9.46 (1.30) | 7.88 (1.40) | 0.354 |
|  | Placebo | 9.07 (1.38) | 5.07 (1.39) |  |
| *Emotion Regulation* |  |  |  |  |
| Reappraisal (ERQ-CA)<sup>49</sup> | GOS | 20.72 (.090) | 21.10 (0.93) | 0.264 |
|  | Placebo | 20.22 (0.86) | 22.62 (0.90) |  |
| Suppression (ERQ-CA)<sup>49</sup> | GOS | 11.57 (0.79) | 11.44 (0.83) | 0.703 |
|  | Placebo | 10.85 (0.76) | 10.11 (0.79) |  |
| TCAQ<sup>50</sup> | GOS | 77.92 (2.54) | 86.33 (2.79) | 0.434 |
|  | Placebo | 73.60 (2.61) | 85.79 (2.63) |  |
| *Sleep* |  |  |  |  |
| PSQI<sup>47</sup> | GOS | 5.43 (0.63) | 4.71 (0.68) | 0.749 |
|  | Placebo | 5.95 (0.64) | 4.81 (0.63) |  |

**Figure 1.**
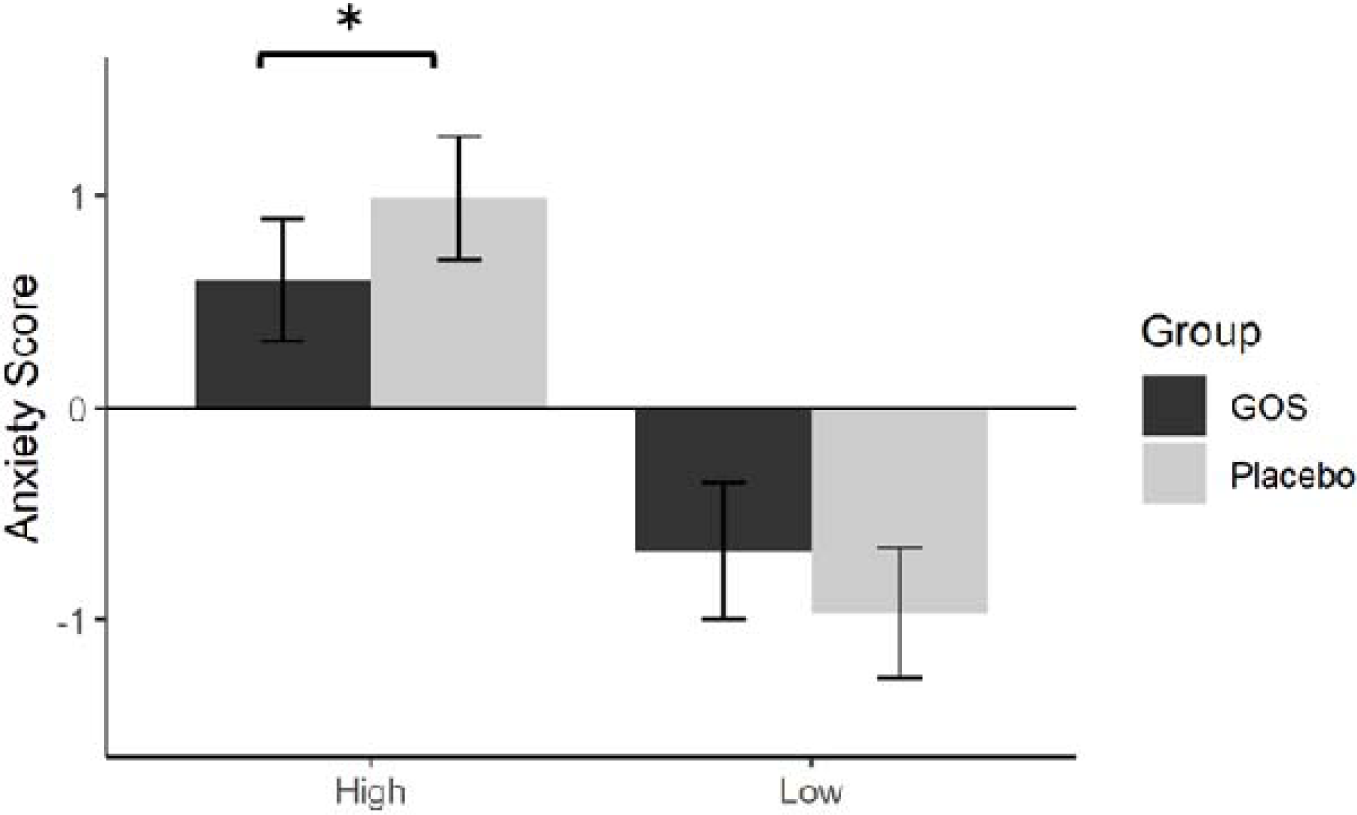
Trait anxiety scores at follow-up (plotted as z-scores, error bars are SE). Scores are reduced in the high anxious (x-axis) GOS group compared to the high anxious placebo group. * p = .062.

### Attentional Dot Probe task

An initial model found no group differences at baseline again confirming suitability of ANCOVA. An interaction of block (masked and unmasked), emotional valence (positive and negative), intervention group (GOS and placebo) and anxiety group (high and low) was found *F*(1,175) = 4.26, *p* = .04, n^2^ = .022. This interaction was then modelled by block separately. There was no significant interaction in the masked block, *F*(1,87) = 0.07, *p* = .788; however, there was a significant interaction with emotional valence, intervention group and anxiety group in the unmasked block, *F*(1,87) = 7.10, *p* = .009, n^2^ = .073. This was further modelled by anxiety grouping independently to examine the influence of intervention group on emotional valance stimuli (**Figure 2**). In the high anxious group, there was a trend towards an interaction between intervention group and valence condition, *F*(1,45) = 3.46, *p* = .070, n^2^ = .071, where participants in the GOS group in comparison to the placebo group showed reduced bias to negative stimuli, GOS *M* = −34.86 ms, *SE* = 18.86 [CI −72.86 – 3.13]; Placebo *M* = 3.67 ms, *SE* = 18.18 [CI −32.95 – 40.29]; and greater bias to positive stimuli, GOS *M* = 3.62 ms, *SE* = 18.90 [CI −34.45 – 41.69]; Placebo *M* = −26.36 ms, *SE* = 18.09 [CI −62.82 – 10.08]. The trend towards interaction between intervention group and valence condition was also found in the low anxious group, *F*(1,41) = 3.49, *p* = .069, n^2^ = .073. Unlike in the high anxious participants, the low anxious placebo group in comparison to the GOS group demonstrated decreased bias to negative stimuli GOS *M* = 2.28 ms, *SE* = 20.07 [CI −38.26 – 42.83]; Placebo *M* = −24.15 ms, *SE* = 19.29 [CI −63.11 – 14.80]; and increased bias to positive stimuli, GOS *M* = −14.63 ms, *SE* = 20.34 [CI −55.73 – 26.45]; Placebo *M* = 31.81 ms, *SE* = 19.33 [CI −63.11 – 14.80].

**Figure 2.**
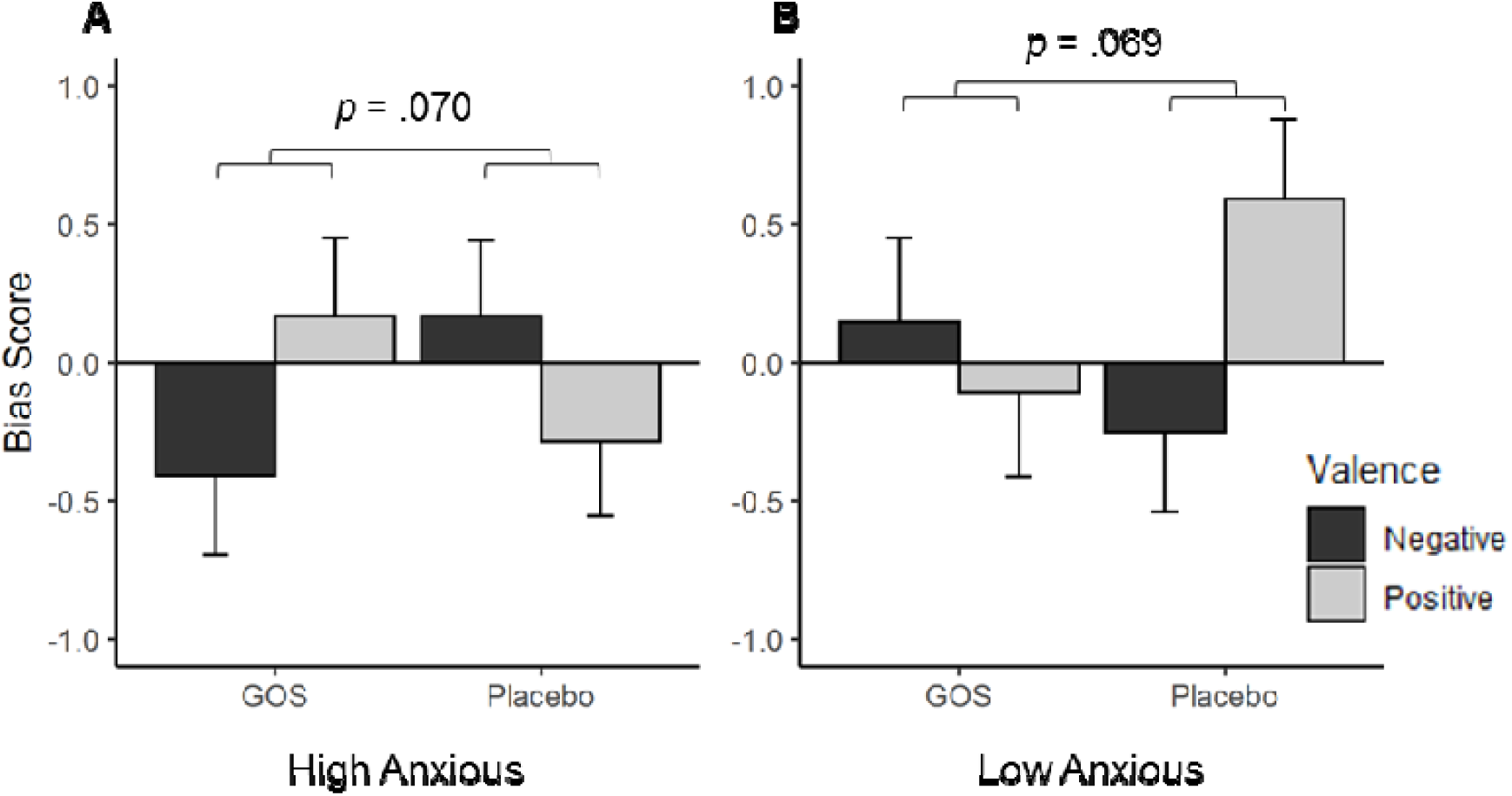
Interaction of attentional vigilance to stimulus valence (y-axis, bias score (z-scores)) by intervention group in the high anxious group, and the low anxious group at follow-up. Error bars illustrate SE **A**, In high anxious group, GOS group participants show a trend towards reduced bias to negative stimuli, and increased bias to positive stimuli in comparison to the placebo group. **B**, in low anxious group the placebo group show a trend towards reduced bias to negative stimuli, and increased bias to positive stimuli in comparison to the GOS group.

### Bacterial composition analysis

Descriptive means of bacterial diversity are displayed in supplementary Table 3. There were no changes in diversity due to intervention or anxiety grouping at follow up as illustrated in the ANCOVA results. There was no main effect of intervention group (*F*(1,43) = 2.08, *p* = .15, n^2^ = .04), anxiety group (*F*(1,43) = 2.63, *p* = .11, n^2^ = .05), or interaction of intervention and anxiety group (*F*(1,43) = 0.01, *p* = .89, n^2^ < .01).

All genera identified in the microbiome were also evaluated using paired non-parametric comparisons within each intervention group from baseline to follow-up, and independent non-parametric comparisons between intervention groups at baseline and follow-up.

#### Within group measures

**Table 2** displays the significant differences from baseline to follow-up for both GOS and placebo groups. Only *Bifidobacterium* was found have significant changes in abundance greater than 1%, and only in the GOS group; baseline, *M* = 6.1, *SE* = 1.6, follow-up, *M* = 9.6, *SE* = 2.0; *p* = 0.032 illustrating a 3.49% increase. There was no change in *Bifidobacterium* abundance in the placebo group; baseline, *M* = 8.2, *SE* = 2.1, follow-up, *M* = 8.0, *SE* = 1.4; *p* = 0.989 (**Figure 3**). Other significant changes across time in genera abundances in the GOS group (supplementary **Figure 1**) were substantially lower; abundances of the bacteria groups unclassified Streptococcaceae, *Falcatimonas, Lachnoanaerobaculum, Lachnospira* decreased in the GOS group, while the abundances of *Faecalicoccus* increased. In the placebo group other changes were observed (**Table 2**) for low abundant members of the phylum of Proteobacteria there was an increase in specific pathogens (e.g. *Escherichia, Salmonella*, and *Shigella*). Notably, the abundances of these groups were not found to be significantly different for subjects in the GOS group (supplementary **Figure 2**).

**Table 2.**
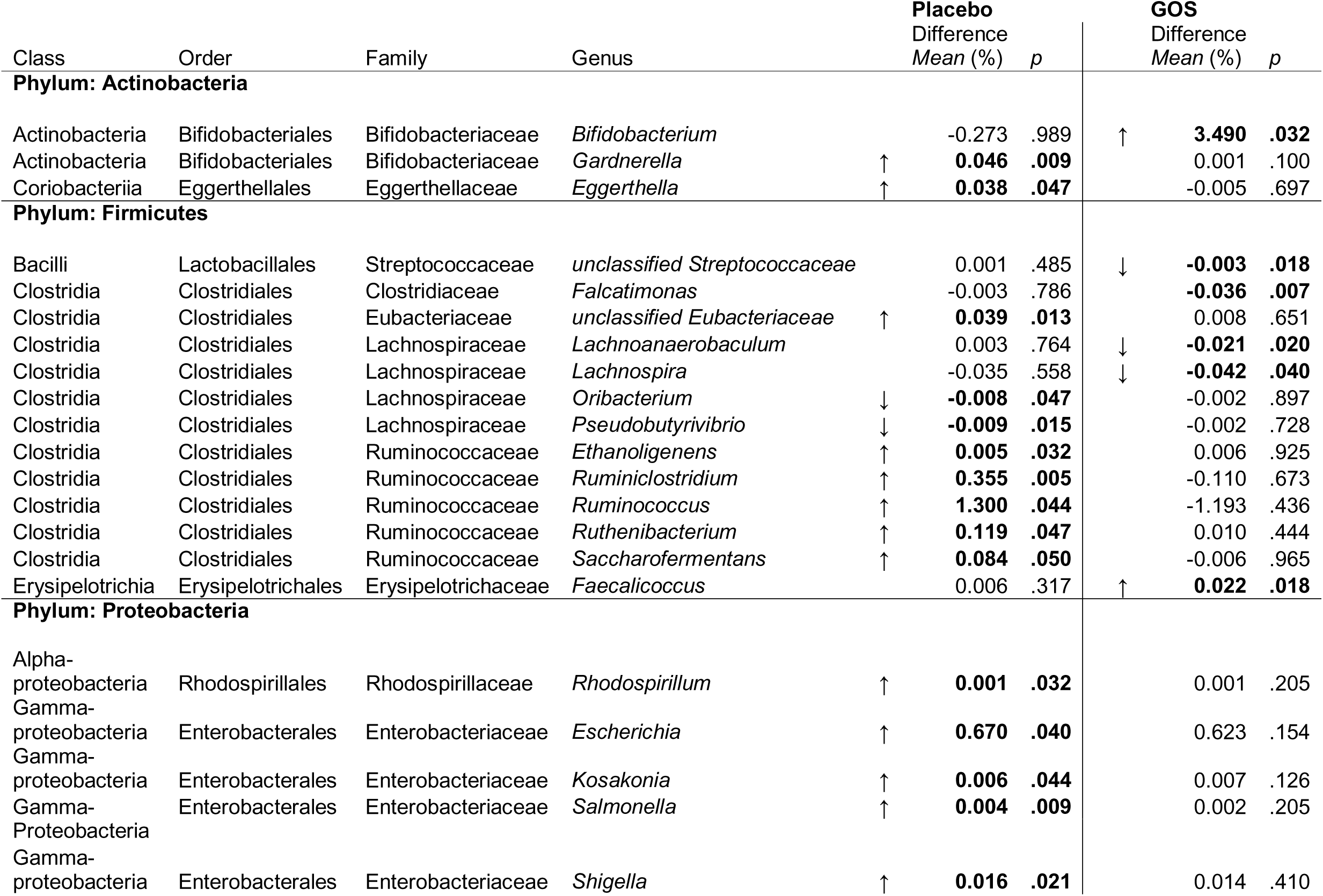
Significant differences in microbial bacteria abundance within GOS and placebo groups from baseline to follow-up. Arrows show direction of effect.

**Figure 3.**
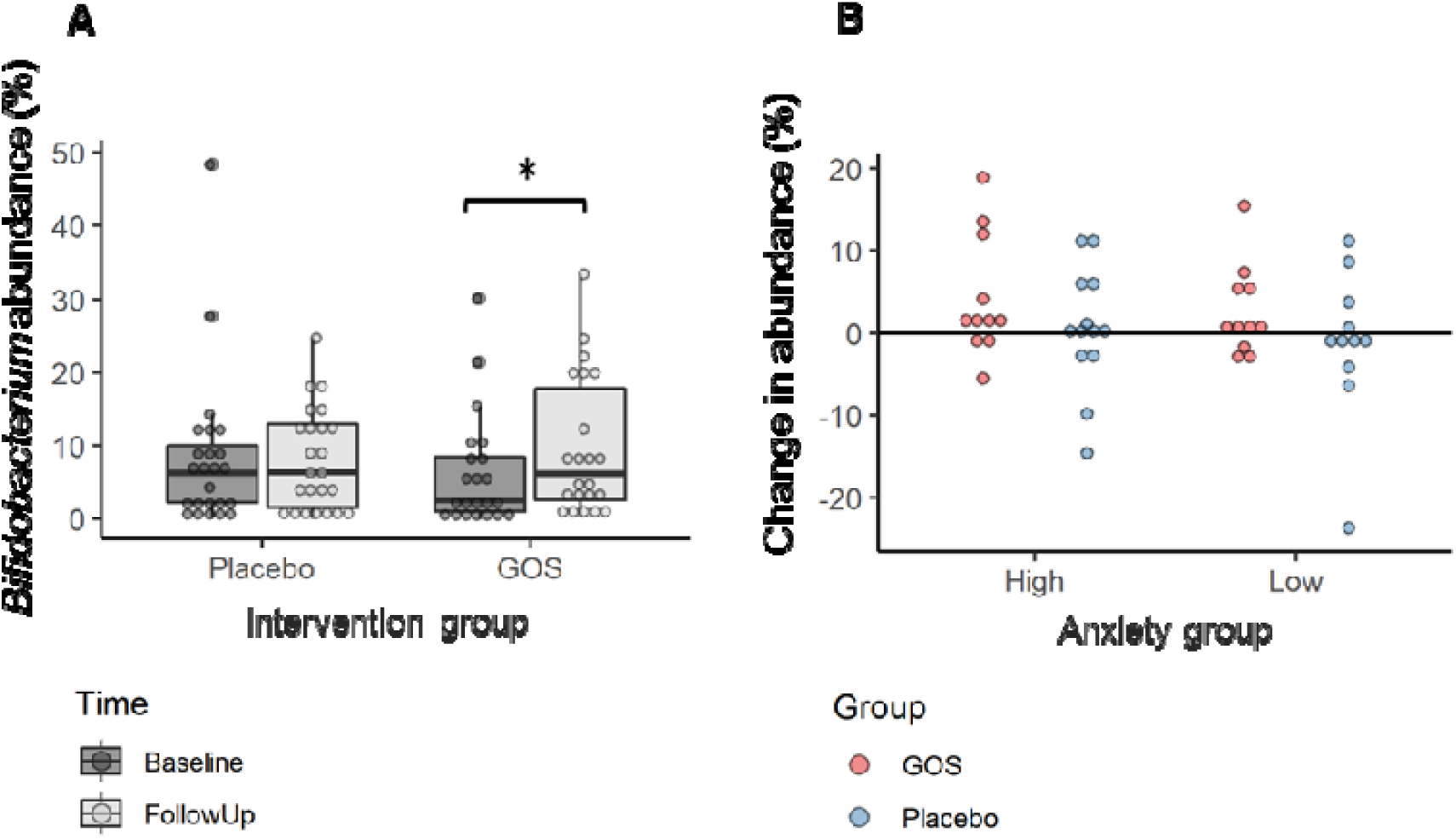
**A** Comparison of *Bifidobacterium* abundance at baseline and follow up for both intervention groups. Boxplots enclose data within the 25^th^-75^th^ percentiles with whiskers a maximum of 1.5 times the distance of box range. Median is indicated by horizontal line, and individual data plotted. There is a significant increase in *Bifidobacterium* in the GOS group at follow up compared to baseline * *p* = .036. **B**. Change in *Bifidobacterium abundance for high and low anxious participants in each intervention group from baseline to follow up. Dots represent individual participants. Those above the zero axis illustrate an increase in the abundance of Bifidobacterium at follow up, those below a decrease*.

#### Between group measures

Between group differences at baseline and follow-up are displayed in **Table 3**, with no genera surpassing a difference in abundance across groups of more than 1%, including *Bifidobacterium* (baseline, *p* = 0.20 or follow-up, *p* = 0.69). At baseline, there were few significant differences between intervention groups (supplementary **Figure 3**). For the genus *Dorea, Catonella, Peptococcus* and *Holdemania* initial abundances were significantly lower in the GOS group in comparison to the placebo group. Bacterial abundances for the genera *Turicibacter* and *Kosakonia* at the baseline samples from the placebo group were lower than in the GOS group. At follow-up, again there are few between group differences, with genera identified in each phylum of Bacteroidetes, Actinobacteria and Firmicutes (supplementary **Figure 4**) showing decreased abundance in the GOS group compared with the placebo group, with little relation easily identified between the specific genera.

**Table 3.** Significant differences between GOS and placebo group at baseline and follow-up. Difference means are calculated in reference to placebo group and arrows show direction of effect.

| Class | Order | Family | Genus | Baseline<br>Difference<br>Mean (%) | *p* |  | Follow-up<br>Difference<br>Mean (%) | *p* |
| --- | --- | --- | --- | --- | --- | --- | --- | --- |
| Phylum: Actinobacteria |  |  |  |  |  |  |  |  |
| Actinobacteria | Actinomycetales | Actinomycetaceae | *Actinomyces* | -0.0001 | 0.106 | ↓ | **0.020** | **0.044** |
| Coriobacteriia | Coriobacteriales | Atopobiaceae | *Atopobium* | -0.002 | 0.484 | ↓ | **0.001** | **0.033** |
| Phylum: Bacteroidetes |  |  |  |  |  |  |  |  |
| Bacteroidia | Bacteroidales | Prevotellaceae | *Prevotella* | 0.765 | 0.912 | ↑ | **-0.271** | **0.019** |
| Sphingobacteriia | Sphingobacteriales | Sphingobacteriaceae | *Sphingobacterium* | -0.003 | 0.79 | ↓ | **0.007** | **0.047** |
| Phylum: Firmicutes |  |  |  |  |  |  |  |  |
| Bacilli | Bacillales | Staphylococcaceae | *Staphylococcus* | 0.002 | 0.435 | ↓ | **0.004** | **0.017** |
| Bacilli | Lactobacillales | Streptococcaceae | unclassified<br>*Streptococcaceae* | -0.001 | 0.355 | ↓ | **0.003** | **0.031** |
| Clostridia | Clostridiales | Clostridiaceae | *Lactonifactor* | 0.002 | 0.766 | ↓ | **0.015** | **0.019** |
| Clostridia | Clostridiales | Clostridiales Family XIII. | *Mogibacterium* | -0.010 | 0.18 | ↓ | **0.018** | **0.007** |
| Clostridia | Clostridiales | Lachnospiraceae | *Dorea* | ↓ **0.335** | **0.028** |  | 0.588 | **0.002** |
| Clostridia | Clostridiales | Lachnospiraceae | *Eisenbergiella* | 0.039 | 0.137 | ↓ | **0.053** | **0.025** |
| Clostridia | Clostridiales | Lachnospiraceae | *Catonella* | ↓ **0.002** | **0.028** |  | 0.001 | 0.68 |
| Clostridia | Clostridiales | Peptococcaceae | *Peptococcus* | ↓ **0.014** | **0.008** |  | 0.005 | 0.234 |
| Erysipelotrichia | Erysipelotrichales | Erysipelotrichaceae | *Holdemania* | ↓ **0.022** | **0.014** |  | 0.004 | 0.434 |
| Erysipelotrichia | Erysipelotrichales | Erysipelotrichaceae | *Turicibacter* | -0.185 | **0.032** |  | -0.002 | 0.344 |
| Phylum: Proteobacteria |  |  |  |  |  |  |  |  |
| Gamma-proteobacteria | Enterobacterales | Enterobacteriaceae | *Kosakonia* | ↑ **-0.006** | **0.028** |  | -0.006 | 0.154 |

### RDA analysis

RDA analysis was performed for both the GOS and placebo group and anxiety group separately contrasting measures across time from baseline to follow-up. Results are displayed in the ordination diagrams in **Figure 4**. For individuals in the GOS group classified as ‘high anxious’ at baseline, genus level microbiota communities showed a trend level difference at follow-up, albeit with a low significance (p = .088). For the GOS low anxious group, there was no significant prediction of microbiome genera on psychological measures across time *p* = .414. In the placebo groups, the low anxious participants illustrate trending effects towards differential microbiota communities from baseline to follow up (*p* = .103) similar to the high anxious GOS participants. There was no such effect in the high anxious participants, *p* = .419.

**Figure 4.**
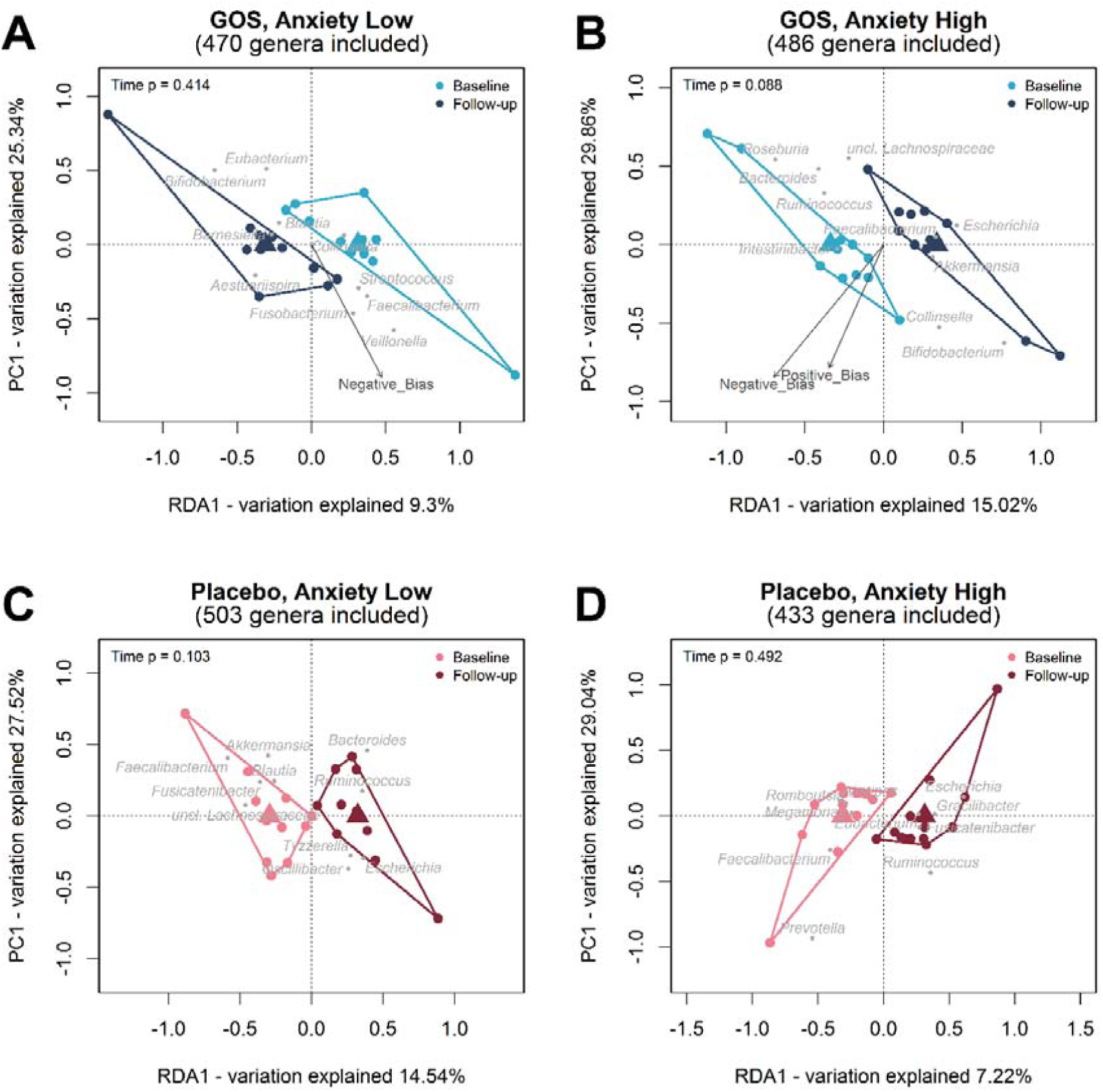
Ordination diagram from RDA for samples from the placebo and GOS group collected at baseline and follow-up, where the RDA indicates the association between Time (explanatory variables (x axis)) and bacterial community data on the Genus level. Scores of the first RDA-axis are plotted on the x-axis and scores of the first PCA-axis are plotted on the y-axis. Individual samples are represented by points that are coloured by time and samples belonging to the same collection time are enveloped. Triangles in the RDA diagrams represent the class centroids. Grey points and labels represent the 10 best-fitting genus level bacterial groups. The psychological measurements (environmental variables) are fitted onto the ordination and indicated by the named arrows. The arrow shows the direction of the (increasing) gradient, and the length of the arrow is proportional to the correlation between the variable and the ordination. Only those psychological measurements with a p value of <0.10 are added to the ordination. The calculated p-value from Unrestricted Permutation Test is added to the upper left corner of the RDA diagram.

## Discussion

In this double-blind placebo-controlled 4-week galacto-oligosaccharides (GOS) supplement intervention study we sought to characterise the influence of GOS on indices of emotional well-being and microbial gut composition in a sample of female participants towards the end of adolescence. We found anxiolytic effects of GOS in high anxious participants in self-reported trait anxiety and trends towards reduced negative emotional bias and increased positive bias in the dot-probe task. Additionally, microbial composition was characterised by increased *Bifidobacterium* abundances at follow up in the GOS group. RDA analysis of microbial composition against psychological measures found trend level separation of bacterial composition in high anxious females at follow up compared to baseline. Altogether, these data indicate that GOS supplementation raises fecal abundances of *Bifidobacterium* and has an anxiolytic influence on emotional wellbeing in high anxious late adolescent females.

The anxiolytic effects evident in this intervention are in line with a prior community study of GOS whereby stress indicators were reduced and emotional behaviour improved following 3 weeks of GOS supplementation^29^, adding corroborative evidence to the literature that GOS supplementation assists in functional enhancement of biological networks underpinning emotion regulation and mood. Anxiolytic and antidepressant effects of multispec pre- and probiotics are apparent in several in-human studies e.g. ^25–27,29,45^, however, results of this study elucidated no demonstrable impact of GOS on measures of mood, depression, emotion regulation or indeed social or state reported anxiety levels. Some *clinical* trials have linked pro- or prebiotic intake to reduced depression^26^ or anxiety^46,47^, others have found no support^48–50^. In these trials, measures of depression and anxiety are usually secondary to primary outcomes in overall improvement in clinical conditions^51^ and are comorbid in the function of existing dysregulated biological systems (e.g. IBS^46,47^, rheumatoid arthritis^52^, fibromyalgia^48^). The finding that GOS only impacts self-reported sub-clinical trait anxiety in this study suggests a dimension of sensitivity in late adolescent females that would benefit from further exploration.

Trait anxiety is a persistent emotional state characterised by doubts, fear and worry. When such a state co-occurs with the final period of maturation in late adolescents, emotion regulation ability is key for determining the trajectory for lifelong behaviours^53^. Animal models have shown that dysbiosis in adolescence results in lasting effects on brain-behaviour interactions^8,15^. While gut-microbiota-brain (GMB) influences are bidirectional, biological systems may be fine-tuned via nutritional intake. This may benefit late adolescence females, especially those subject to transient suboptimal emotional regulation skills in the final stages of maturation (e.g. displaying symptoms of stress and anxiety). Nutritional supplementation and consequential influence on GMB may prove effective in stabilising some of these symptoms in this age group. Probiotics are known to influence the gut-brain axis via endocrine, immune and neural pathways^27,54,55^. For example, *Lactobacillus rhamnosus* intake exerts influence on GABA receptors in the brain^54^, which cooperates with excitatory glutamate to regulate brain function. Glutamate and GABA are respectively excitatory and inhibitory neurotransmitters that are essential in typical cognitive development^56^. In anxious individuals, disruption to the balance of this relationship is correlated with poorer cognitive control^57^ and influences emotion regulation ability. The intake of pre- and probiotics may contribute to the harmonisation of the excitatory/inhibitory balance.

We affirmed a clear influence of GOS intake on *Bifidobacterium* increase across time in comparison to placebo group. Data are also suggestive of a decrease in potential pathogens. While measures of microbiota diversity are similar across groups and time, it may be that GOS intake resulted in the prevention of potentially unhealthy bacteria, offering protective benefits. Admittedly, it is difficult to disentangle if the influence of GOS observed in these data is due to the stimulation of *Bifidobacterium* growth, or the reduction in proteobacteria. Naturally, diet is likely a key component of this relationship and in this sample, we found little support for dietary changes in groups across time, excepting reduced sugar intake in the GOS group, or between groups. This strengthens the hypothesis that GOS exerts beneficial influence on well-being, yet the mechanisms of this require further investigation. Regardless, the subtle fine tuning of microbiota with GOS intake may be enough to assist in the biological regulation of emotional pathways and contribute to improved well-being in pre-clinical populations.

Of note in the outcome of this study is the relatively small effect sizes and trend level p-values of our self-report psychological measures and dot probe task. The dot-probe task encapsulates attentional biases to emotional stimuli. Prevalent, heightened anxiety commonly results in attentional bias to negative, or threating information^30^, and can be reduced by attentional bias modification training^58^. Attentional training works to modify emotion regulation networks in the brain^59^ however, there are questions surrounding the consistency of the manifestation of attentional biases at the individual level^59^. In the absence of attentional bias training, results from the dot probe task were indicative of trends towards reduced negative bias and increased positive bias to emotional stimuli in the high anxious GOS group, a pattern that was also observed in the low-anxious placebo group. While this may indicate that high anxious GOS participants have undergone alterations in cognitive-emotional processing compared to the high anxious placebo group, it is difficult to cleanly interpret as the low anxious GOS group results do not mirror their placebo counterparts. Although there are trends towards interventional effects, individual variability in anxiety expression within intervention groups may be influenced by variables not accounted for in this study.

Similar community sample research produce equally small effect sizes on comparative measures^29^. This might be anticipated in an intervention in sub-clinical typical populations examining factors at group level where specific pathways and mechanisms are yet to be fully established. Well defined operational measures with strong correlations to mediating pathways as exhibited in animal studies may result in more targeted therapeutic potential of GOS in humans. Herein, GOS has been established to increase *Bifidobacterium* abundance in 4 weeks in co-occurrence with reduced anxiety manifestation, with indications of modification of attentional bias in a sample of adolescent females. Data presented here are indicative that GOS may be effective in influencing the expression of anxiety and would benefit from further research specifying potential pathways of this effect.

## Data Availability

Data and scripts for reproducing analysis in this manuscript are freely available.

https://osf.io/ngmsu/

## Acknowledgements

This research was supported by faculty research fund from the Faculty of Health and Medical Sciences, University of Surrey, UK to KCK. FrieslandCampina provided the galacto-oligosaccharides (GOS, prebiotics) used in this study.

## Contributions

KCK conceived the study with input from PWJB, PS and KH. CM, OB and NJ acquired the data. NJ and BvdB analysed and interpreted the data. All authors have drafted or substantially revised the article and have approved the present version. All authors agree to be personally accountable in matters relating to their own contributions.

## Data availability

Data and code for reproducible analysis are available at DOI 10.17605/OSF.IO/NGMSU

## Supplementary material

**Supp. Table 1.**
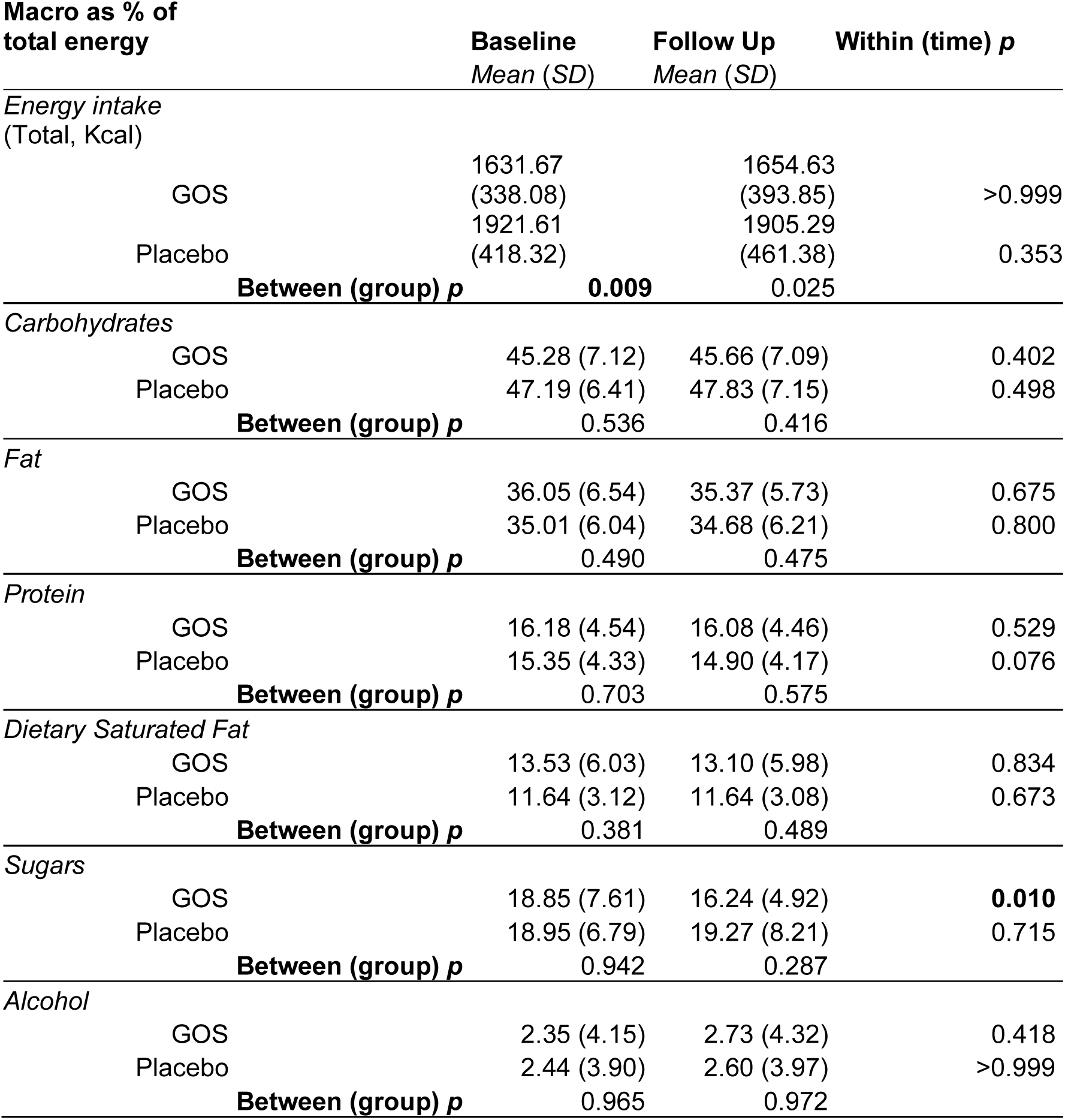
Means and non-parametric test significance values of food diary entries for the first and final four days of supplement intake.

| **Macro as % of**<br>**total energy** | **Baseline**<br>**Mean (SD)** | **Follow Up**<br>**Mean (SD)** | **Within (time) p** |
| --- | --- | --- | --- |
| *Energy intake*<br>*(Total, Kcal)* |  |  |  |
| GOS | 1631.67<br>(338.08) | 1654.63<br>(393.85) | >0.999 |
| Placebo | 1921.61<br>(418.32) | 1905.29<br>(461.38) | 0.353 |
| **Between (group) p** | **0.009** | 0.025 |  |
| *Carbohydrates* |  |  |  |
| GOS | 45.28 (7.12) | 45.66 (7.09) | 0.402 |
| Placebo | 47.19 (6.41) | 47.83 (7.15) | 0.498 |
| **Between (group) p** | 0.536 | 0.416 |  |
| *Fat* |  |  |  |
| GOS | 36.05 (6.54) | 35.37 (5.73) | 0.675 |
| Placebo | 35.01 (6.04) | 34.68 (6.21) | 0.800 |
| **Between (group) p** | 0.490 | 0.475 |  |
| *Protein* |  |  |  |
| GOS | 16.18 (4.54) | 16.08 (4.46) | 0.529 |
| Placebo | 15.35 (4.33) | 14.90 (4.17) | 0.076 |
| **Between (group) p** | 0.703 | 0.575 |  |
| *Dietary Saturated Fat* |  |  |  |
| GOS | 13.53 (6.03) | 13.10 (5.98) | 0.834 |
| Placebo | 11.64 (3.12) | 11.64 (3.08) | 0.673 |
| **Between (group) p** | 0.381 | 0.489 |  |
| *Sugars* |  |  |  |
| GOS | 18.85 (7.61) | 16.24 (4.92) | **0.010** |
| Placebo | 18.95 (6.79) | 19.27 (8.21) | 0.715 |
| **Between (group) p** | 0.942 | 0.287 |  |
| *Alcohol* |  |  |  |
| GOS | 2.35 (4.15) | 2.73 (4.32) | 0.418 |
| Placebo | 2.44 (3.90) | 2.60 (3.97) | >0.999 |
| **Between (group) p** | 0.965 | 0.972 |  |

**Supp. Table 2.**
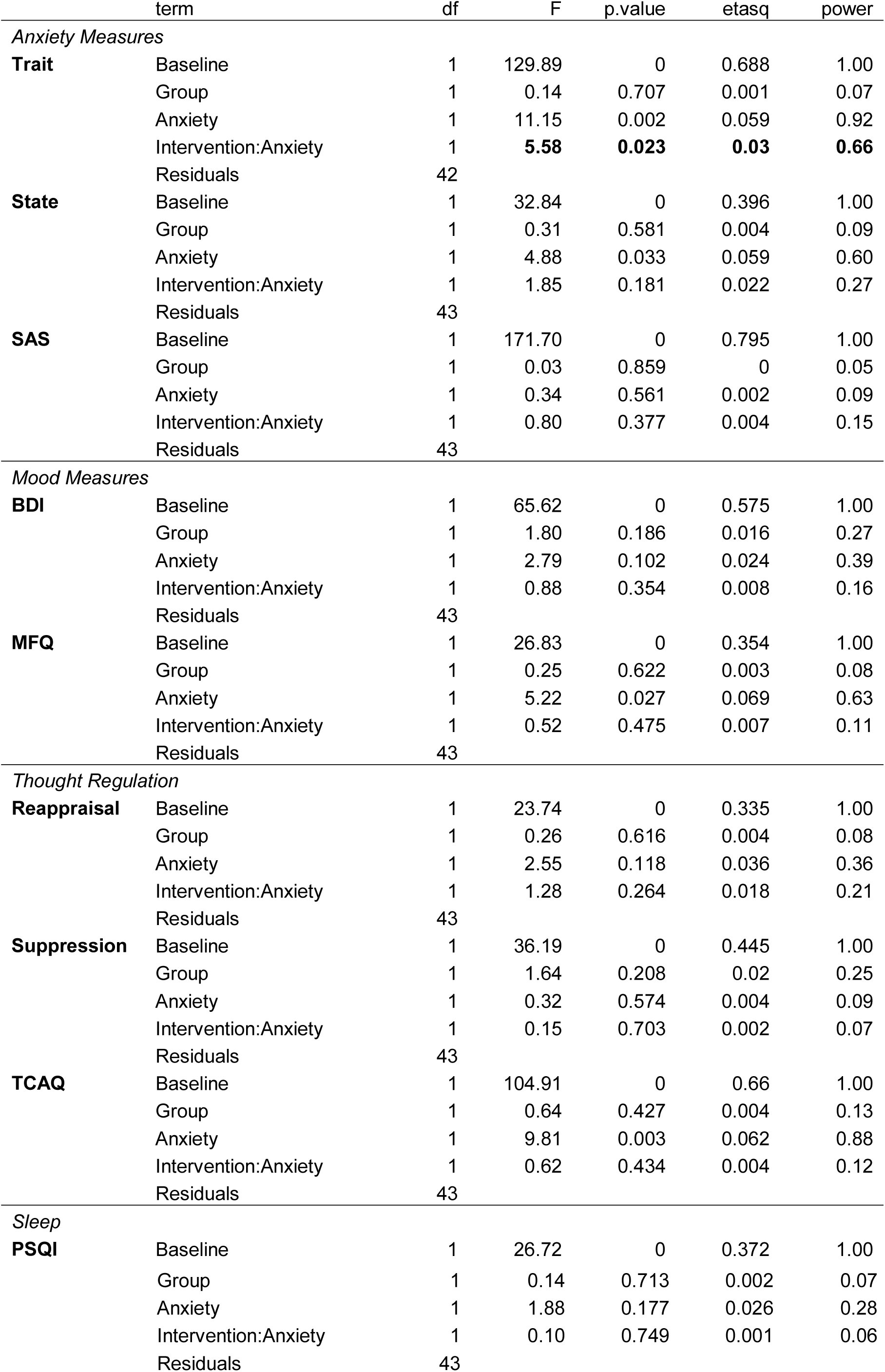
ANCOVA interaction for SRMs

**Supp. Table 3.** Descriptive statistics for bacterial diversity by intervention and anxiety group.

|  |  | Baseline<br>*Mean (SD)* | Follow up<br>*Mean (SD)* |
| --- | --- | --- | --- |
| ***Anxiety*** | ***Intervention*** |  |  |
| High | GOS | 4.62 (1.03) | 4.65 (0.93) |
|  | Placebo | 4.76 (0.61) | 5.08 (0.85) |
| Low | GOS | 4.99 (0.68) | 4.95 (0.66) |
|  | Placebo | 4.93 (0.58) | 5.28 (0.59) |

**Supp. Figure 1.**
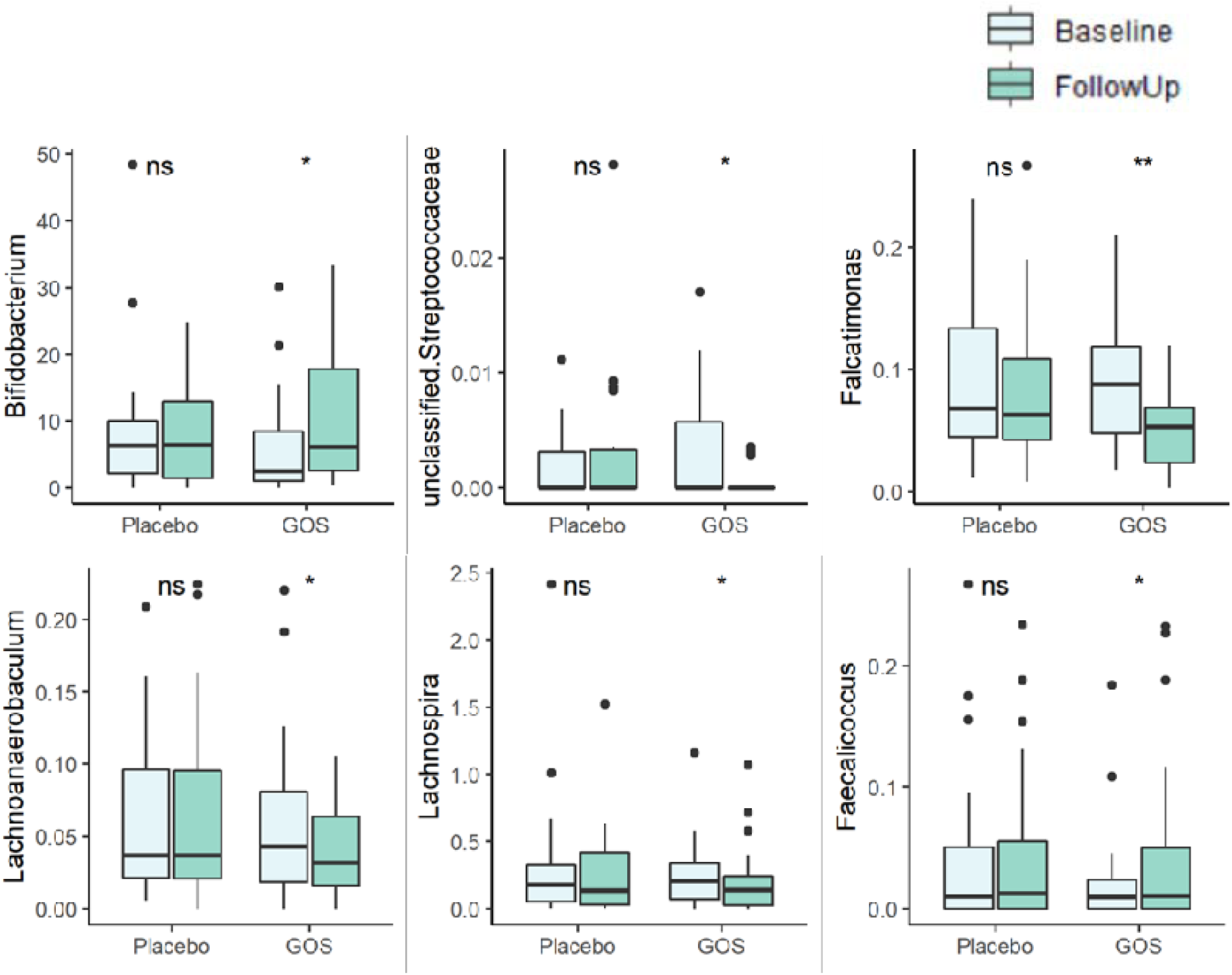
*Differences found in genera in GOS group from baseline to follow-up, with placebo group included for comparison.* Boxplots enclose data within the 25^th^-75^th^ percentiles with whiskers a maximum of 1.5 times the distance of box range. Median is indicated by horizontal line, and outlying data points plotted.

**Supp. Figure 2.**
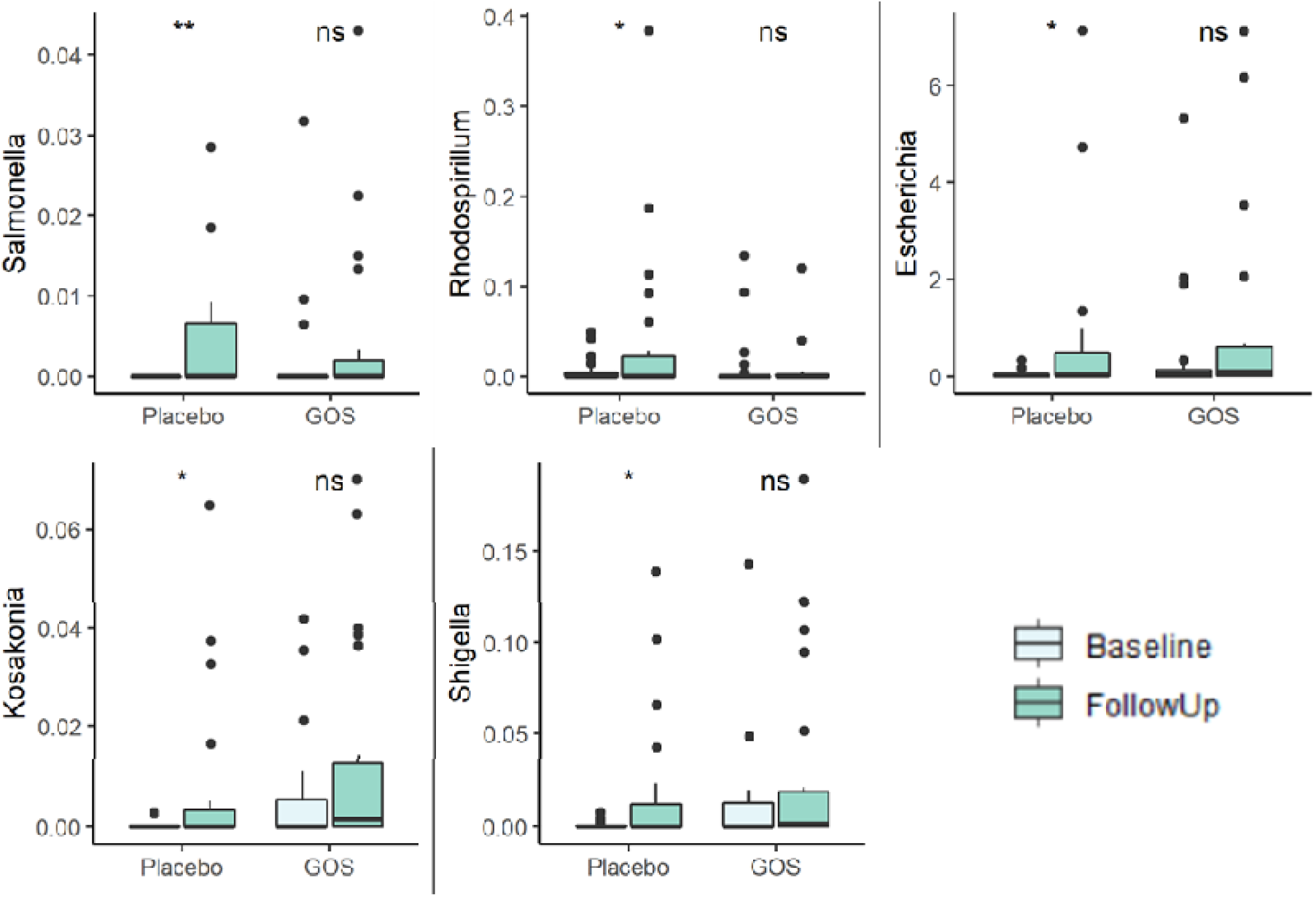
*Proteobacteria genera which significantly increased in the placebo group from baseline to follow-up. GOS group data is shown for comparison. While these bacteria are also present in the GOS group, the increase at follow-up is not as substantial as that in the placebo group.* Boxplots enclose data within the 25^th^-75^th^ percentiles with whiskers a maximum of 1.5 times the distance of box range. Median is indicated by horizontal line, and outlying data points plotted.

**Supp. Figure 3.**
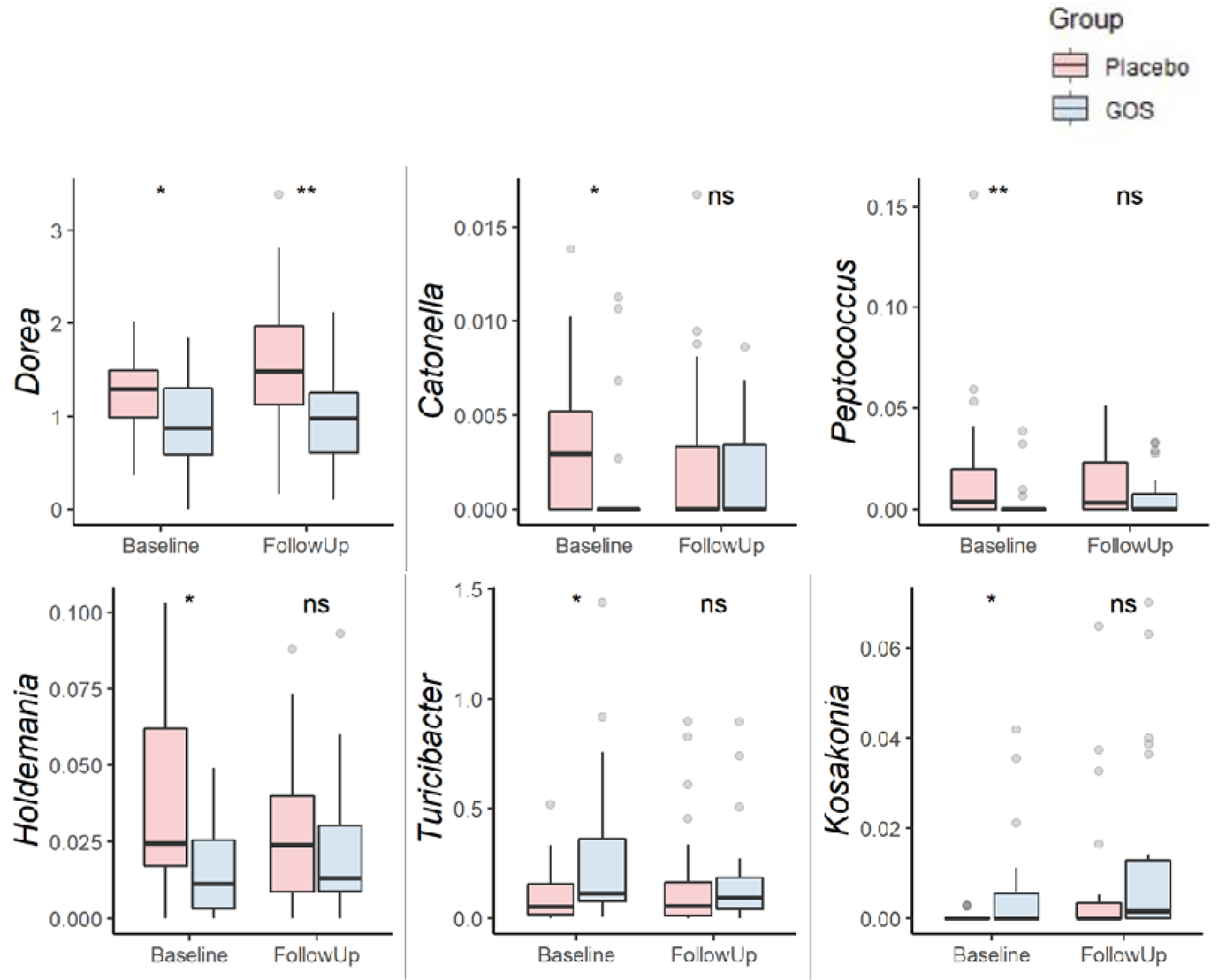
*Baseline differences in genera between groups, with follow-up data included for comparison.* Boxplots enclose data within the 25^th^-75^th^ percentiles with whiskers a maximum of 1.5 times the distance of box range. Median is indicated by horizontal line, and outlying data points plotted.

**Supp. Figure 4.**
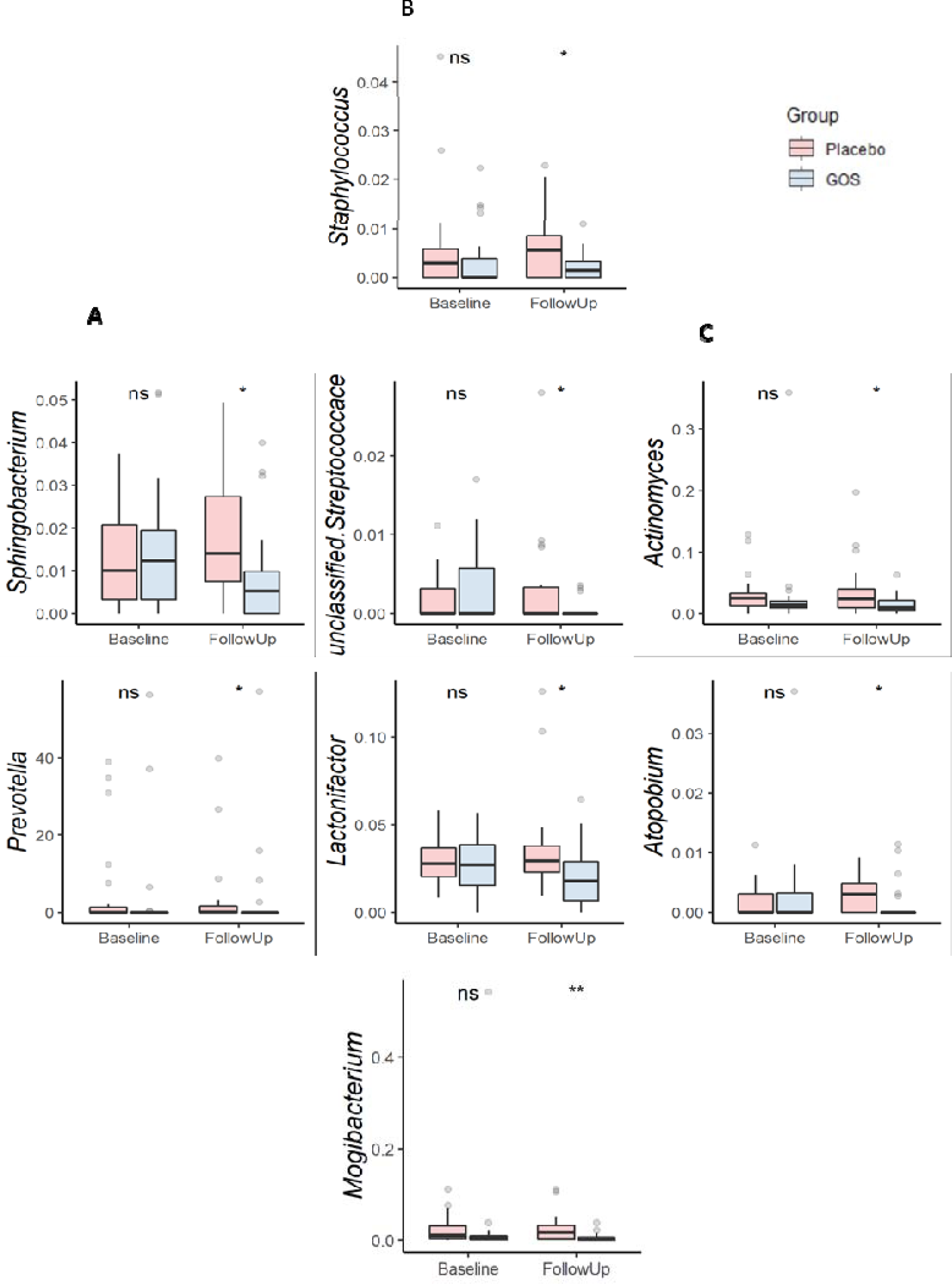
*Intervention group differences at follow-up. Column **A**. Bacteroidetes genera Prevotella and Sphingobacterium are decreased for the GOS group compared to the placebo group. Column **B**. Firmicutes genera again all reduced in the GOS group compared to the placebo group, and column **C**. Actinobacteria genera Actinomyces and Atopobium similarly are reduced in the GOS compared to the placebo intervention group.* Boxplots enclose data within the 25^th^-75^th^ percentiles with whiskers a maximum of 1.5 times the distance of box range. Median is indicated by horizontal line, and outlying data points plotted.

## References

1. Tang, F., Reddy, B. L. & Saier Jr., M. H. Psychobiotics and Their Involvement in Mental Health. J. Mol. Microbiol. Biotechnol. 24, 211–214 (2014).

2. Sarkar, A. et al. Psychobiotics and the Manipulation of Bacteria-Gut-Brain Signals. Trends Neurosci. 39, 763–781 (2016).

3. McVey Neufeld, K.-A., Luczynski, P., Seira Oriach, C., Dinan, T. G. & Cryan, J. F. What’s bugging your teen?—The microbiota and adolescent mental health. Neurosci. Biobehav. Rev. 70, 300–312 (2016).

4. Consortium, T. H. M. P. et al. A framework for human microbiome research. Nature 486, 215–221 (2012).

5. Grenham, S., Clarke, G., Cryan, J. F. & Dinan, T. G. Brain-gut-microbe communication in health and disease. Front. Physiol. 2 DEC, 94 (2011).

6. Grossman, M. I. Neural and Hormonal Regulation of Gastrointestinal Function: An Overview. Annu. Rev. Physiol. 41, 27–27 (2003).

7. Mayer, E. A., Knight, R., Mazmanian, S. K., Cryan, J. F. & Tillisch, K. Gut Microbes and the Brain: Paradigm Shift in Neuroscience. J. Neurosci. 34, 15490–15496 (2014).

8. Turnbaugh, P. J. et al. The effect of diet on the human gut microbiome: A metagenomic analysis in humanized gnotobiotic mice. Sci. Transl. Med. 1, 6ra14–6ra14 (2009).

9. Oldham, M. C. et al. Functional organisation of the transcriptome in human brain. 11, (2008).

10. Desbonnet, L., Garrett, L., Clarke, G., Bienenstock, J. & Dinan, T. G. The probiotic Bifidobacteria infantis: An assessment of potential antidepressant properties in the rat. J. Psychiatr. Res. 43, 164–174 (2008).

11. Foster, J. A. & McVey Neufeld, K.-A. Gut–brain axis: how the microbiome influences anxiety and depression. Trends Neurosci. 36, 305–312 (2013).

12. Lloyd-Price, J. et al. Strains, functions and dynamics in the expanded Human Microbiome Project. Nature 550, 61–66 (2017).

13. Mccartney, A. L., Parracho, H. M. R. T., Bingham, M. O. & Gibson, G. R. Differences between the gut microflora of children with autistic spectrum disorders and that of healthy children. J. Med. Microbiol. 54, 987–991 (2005).

14. Mayer, E. A. Gut feelings: the emerging biology of gut–brain communication. Nat. Rev. Neurosci. 12, 453–466 (2011).

15. Stilling, R. M. et al. Microbes & neurodevelopment – Absence of microbiota during early life increases activity-related transcriptional pathways in the amygdala. Brain. Behav. Immun. 50, 209–220 (2015).

16. Chen, J. et al. Effects of gut microbiota on the microRNA and mRNA expression in the hippocampus of mice. Behav. Brain Res. 322, 34–41 (2017).

17. Diaz Heijtz, R. et al. Normal gut microbiota modulates brain development and behavior. Proc. Natl. Acad. Sci. U. S. A. 108, 3047–52 (2011).

18. David, L. A. et al. Diet rapidly and reproducibly alters the human gut microbiome. Nature 505, 559–63 (2014).

19. Bindels, L. B., Delzenne, N. M., Cani, P. D. & Walter, J. Towards a more comprehensive concept for prebiotics. Nat. Rev. Gastroenterol. Hepatol. 12, 303–310 (2015).

20. Dinan, T. G., Stanton, C. & Cryan, J. F. Psychobiotics: a novel class of psychotropic. Biol. Psychiatry 74, 720–6 (2013).

21. Burnet, P. W. J. & Cowen, P. J. Psychobiotics highlight the pathways to happiness. Biol. Psychiatry 74, 708–9 (2013).

22. Boehm, G. et al. Prebiotics in Infant Formulas. J. Clin. Gastroenterol. 38, S76–S79 (2004).

23. Gibson, G. R. et al. Dietary prebiotics: current status and new definition. (2010).

24. Barile, D. & Rastall, R. A. Human milk and related oligosaccharides as prebiotics. Current Opinion in Biotechnology 24, 214–219 (2013).

25. Messaoudi, M. et al. Beneficial psychological effects of a probiotic formulation (Lactobacillus helveticus R0052 and Bifidobacterium longum R0175) in healthy human volunteers. Gut Microbes 2, 256–261 (2011).

26. Steenbergen, L., Sellaro, R., van Hemert, S., Bosch, J. A. & Colzato, L. S. A randomized controlled trial to test the effect of multispecies probiotics on cognitive reactivity to sad mood. Brain. Behav. Immun. 48, 258–264 (2015).

27. Tillisch, K. et al. Consumption of Fermented Milk Product With Probiotic Modulates Brain Activity. Gastroenterology 144, 1394-1401.e4 (2013).

28. Zmora, N. et al. Personalized Gut Mucosal Colonization Resistance to Empiric Probiotics Is Associated with Unique Host and Microbiome Features. Cell 174, 1388-1405.e21 (2018).

29. Schmidt, K. et al. Prebiotic intake reduces the waking cortisol response and alters emotional bias in healthy volunteers. Psychopharmacology (Berl). 232, 1793–801 (2015).

30. Bar-Haim, Y., Lamy, D., Pergamin, L., Bakermans-Kranenburg, M. J. & van IJzendoorn, M. H. Threat-related attentional bias in anxious and nonanxious individuals: A meta-analytic study. Psychol. Bull. 133, 1–24 (2007).

31. Kessler, R. C. et al. Lifetime prevalence and age-of-onset distributions of DSM-IV disorders in the national comorbidity survey replication. Archives of General Psychiatry 62, 593–602 (2005).

32. Trentacosta, C. J. & Fine, S. E. Emotion Knowledge, Social Competence, and Behavior Problems in Childhood and Adolescence: A Meta-analytic Review. Soc. Dev. 19, 1–29 (2010).

33. Spielberger, C. D., Gorsuch, R. L., Lushene, R., Vagg, P. R. & Jacobs, G. A. State-Trait Anxiety Inventory for Adults. (Mind Garden Inc., 1983).

34. La Greca, A. M. Manual for the Social Anxiety Scales for Children and Adolescents - Revised. (University of Miami, 1999).

35. Beck, A. T., Steer, R. A. & Brown, G. K. BDI-II: Beck Depression Inventory Manual. (Psychological Corporation, 1996).

36. Angold, A. et al. Development of a short questionnaire for use in epidemiological studies of depression in children and asolescents. J. Methods Psychiatr. Res. 5, 237–249 (1995).

37. Gullone, E. & Taffe, J. The Emotion Regulation Questionnaire for Children and Adolescents (ERQ–CA): A psychometric evaluation. Psychol. Assess. 24, 409–417 (2012).

38. Luciano, J. V., Algarabel, S., Tomás, J. M. & Martínez, J. L. Development and validation of the thought control ability questionnaire. Pers. Individ. Dif. 38, 997–1008 (2004).

39. Buysse, D. J., Reynolds, C. F., Monk, T. H., Berman, S. R. & Kupfer, D. J. The Pittsburgh sleep quality index: A new instrument for psychiatric practice and research. Psychiatry Res. 28, 193–213 (1989).

40. Wechsler, D. Wechsler Abbreviated Scale of Intelligence. (The Psychological Corporation, 1999).

41. Edgar, R. C. Search and clustering orders of magnitude faster than BLAST. Bioinformatics 26, 2460–2461 (2010).

42. Wang, Q., Garrity, G. M., Tiedje, J. M. & Cole, J. R. Naive Bayesian Classifier for Rapid Assignment of rRNA Sequences into the New Bacterial Taxonomy. Appl. Environ. Microbiol. 73, 5261–5267 (2007).

43. Oksanen, J. et al. vegan: Community Ecology Package. (2019).

44. Team, R. C. R: A language and environment for statistical computing. R Foundation for Statistical Computing. (2018).

45. Luo, J. et al. Ingestion of Lactobacillus strain reduces anxiety and improves cognitive function in the hyperammonemia rat. Sci. China Life Sci. 57, 327–335 (2014).

46. Silk, D. B. A., Davis, A., Vulevic, J., Tzortzis, G. & Gibson, G. R. Clinical trial: the effects of a trans-galactooligosaccharide prebiotic on faecal microbiota and symptoms in irritable bowel syndrome. Aliment. Pharmacol. Ther. 29, 508–518 (2009).

47. Azpiroz, F. et al. Effects of scFOS on the composition of fecal microbiota and anxiety in patients with irritable bowel syndrome: a randomized, double blind, placebo controlled study. Neurogastroenterol. Motil. 29, e12911 (2017).

48. Roman, P. et al. A Pilot Randomized Controlled Trial to Explore Cognitive and Emotional Effects of Probiotics in Fibromyalgia. Sci. Rep. 8, 10965 (2018).

49. Reale, M. et al. Daily intake of Lactobacillus casei Shirota increases natural killer cell activity in smokers. Br. J. Nutr. 108, 308–314 (2012).

50. Sanchez, M. et al. Effects of a Diet-Based Weight-Reducing Program with Probiotic Supplementation on Satiety Efficiency, Eating Behaviour Traits, and Psychosocial Behaviours in Obese Individuals. Nutrients 9, 284 (2017).

51. Liu, X., Cao, S. & Zhang, X. Modulation of Gut Microbiota–Brain Axis by Probiotics, Prebiotics, and Diet. J. Agric. Food Chem. 63, 7885–7895 (2015).

52. Vaghef-Mehrabany, E. et al. Probiotic supplementation improves inflammatory status in patients with rheumatoid arthritis. Nutrition 30, 430–435 (2014).

53. Haller, S. P. W., Cohen Kadosh, K., Scerif, G. & Lau, J. Y. F. Social anxiety disorder in adolescence: How developmental cognitive neuroscience findings may shape understanding and interventions for psychopathology. Developmental Cognitive Neuroscience 13, 11–20 (2015).

54. Bravo, J. A. et al. Ingestion of Lactobacillus strain regulates emotional behavior and central GABA receptor expression in a mouse via the vagus nerve. Proc. Natl. Acad. Sci. U. S. A. 108, 16050–5 (2011).

55. Collins, S. & Reid, G. Distant Site Effects of Ingested Prebiotics. Nutrients 8, (2016).

56. Luján, R., Shigemoto, R. & López-Bendito, G. Glutamate and GABA receptor signalling in the developing brain. Neuroscience 130, 567–580 (2005).

57. Morgenroth, E. et al. Altered relationship between prefrontal glutamate and activation during cognitive control in people with high trait anxiety. Cortex 117, 53–63 (2019).

58. Hakamata, Y. et al. Attention bias modification treatment: a meta-analysis toward the establishment of novel treatment for anxiety. Biol. Psychiatry 68, 982–90 (2010).

59. Ghassemzadeh, H., Rothbart, M. K. & Posner, M. I. Anxiety and Brain Networks of Attentional Control. Cogn. Behav. Neurol. 32, 54–62 (2019).

